# Optimizing culture-free approaches to recover high quality *M. tuberculosis* genomic variation

**DOI:** 10.64898/2025.12.16.25342406

**Authors:** Katharine S. Walter, Paulo César Pereira Dos Santos, Allison Carey, Salika M. Shakir, Caroline Colijn, Ted Cohen, Barun Mathema, Julio Croda, Jason R. Andrews

## Abstract

**Background:** *Mycobacterium tuberculosis* (*Mtb*) genomic epidemiology often relies on culturing patient sputum, a time and labor-intensive process. Hybrid capture approaches have been successfully used to enrich *Mtb* DNA from complex clinical samples, yet the accuracy of variant identification from captured samples has not been systematically evaluated.

**Methods:** We created artificial strain mixtures of two well-characterized *Mtb* isolates such that the minor strain comprised 0-50% of *Mtb* DNA and serially diluted the *Mtb* DNA into human DNA to simulate diagnostic samples with different sputum smear burdens (32 samples). We also prospectively collected paired *Mtb* diagnostic cultures and sputum submitted to a national diagnostic laboratory (7 sample pairs). We performed hybrid capture and Illumina whole genome sequencing for all samples. For the artificial strain mixtures, we measured hybrid capture efficiency, the percentage of total reads mapping to *Mtb*, and performance of fixed and minority variant identification. For the diagnostic samples, we compared the number, identity of, and minor allele frequencies of minority variants identified in the cultured and hybrid captured samples.

**Results:** In the artificial strain mixture experiment, hybrid capture efficiency was 97% when *Mtb* comprised 0.01% of the input DNA. Single nucleotide polymorphism (SNP) identification via hybrid capture had a sensitivity ≥ 91% and precision ≥ 97% for *Mtb* lineages 4.1.2.1 and 4.9, excluding PE/PPE genes, when *Mtb* comprised 0.01% of input DNA. Observed minor allele frequencies were closely correlated (r=0.60 to r=0.79, p < 0.001) with input minor allele frequences across all dilutions. Among paired diagnostic samples, hybrid capture efficiency was high, 95%. However, four of the seven captured sputa samples were overwhelmed with *Pseudomonas* contamination, which comprised >25% of sequence reads. We did not detect a significant difference in the number of minority variants identified in cultured (median: 14, range: 9-23) and hybrid captured samples (median: 22 variants, range 4-328, p=0.2) and minor allele frequences were correlated (r = 0.95, p < 0.001)

**Conclusions:** Hybrid capture of diagnostic sputa samples efficiently generates accurate *Mtb* whole genome sequences and minority variant calls. Hybrid capture may offer an alternative to culture-based sequencing that could extend the coverage of genomic epidemiology studies.

**Impact Statement:** *Mycobacterium tuberculosis* (*Mtb*) genomic epidemiology often relies on culturing patient sputa, which is time and labor-intensive. Hybrid capture approaches enrich *Mtb* DNA from complex clinical samples, yet the efficiency and accuracy of identification of both consensus and minority variants from captured samples has not been systematically evaluated. To address these questions, we assessed the accuracy of hybrid capture both in experimental strain mixtures of well-characterized *Mtb* isolates and in clinical diagnostic samples. We found that hybrid capture of sputa samples can be used to efficiently sequence *Mtb* DNA from complex mixtures and that variant identification is highly accurate. Our results suggest that hybrid capture may offer an alternative to culture-based sequencing that could extend the coverage of genomic epidemiology studies.

## Introduction

Genomic epidemiology is a powerful tool for understanding *Mycobacterium tuberculosis* (*Mtb*) transmission patterns, evolutionary processes, and the mechanisms of antibiotic resistance.^1^ Current routine approaches for *Mtb* genomic epidemiology rely on culturing patient sputum to generate sufficient *Mtb* DNA for sequencing.^1^ Mycobacterial culture is time and labor-intensive and requires laboratory biosafety level 3 infrastructure. Culture may also impose a bottleneck on *Mtb* variation, as *Mtb* diversity present in sputa may be lost or outcompeted in culture, and some strains are not successfully isolated in culture.^2–4^ However, a recent study has reported no loss of *Mtb* variation attributable to bacterial culture.^5^ Further, producing sputum is difficult for many patients, including children, HIV-positive individuals, and asymptomatic individuals.^6^ While other clinical specimen types can be used for culture, this is not routinely done, with the result that many individuals are excluded from *Mtb* genomic studies.

Several approaches have been proposed to bypass culture and enrich *Mtb* DNA or deplete human DNA, enabling direct sequencing from clinical samples. This is necessary because *Mtb* DNA often comprises a small fraction of the total DNA.^7,8^ Metagenomic or shotgun sequencing—in which total sample DNA is sequenced—generates sequence reads that are primarily non *Mtb*-derived and do not result in sufficient coverage for phylogenetic and transmission inference nor genotyping of antimicrobial resistance.^7^ Several approaches for eukaryotic DNA depletion, often with detergents, have increased the efficiency of metagenomic sequencing of *Mycobacterium abscessus* and other respiratory pathogens.^9,10^ Hybrid capture, the selective enrichment of target DNA via hybridization with complementary RNA probes, has enabled the *M. tuberculosis* sequencing directly from sputum samples.^8,11–13^ More recently, a tiled amplicon sequencing approach, based on the widely used approach for sequencing SARS-CoV-2 and other viruses, has been adapted for *Mtb*.^14^

Previous studies have demonstrated that hybrid capture enables sequencing of *Mtb* DNA from complex clinical samples, enabling transmission inference and genotypic prediction of antimicrobial resistance ^8,11–13^ Hybrid capture has also used successfully to sequence *Mtb* DNA from environmental samples.^15^ Yet the accuracy of variant identification from hybrid captured samples has not been systematically evaluated, and whether variant accuracy varies with *Mtb* genomic background is unknown. Further, whether there is a limit of detection, a minimum sample proportion of *Mtb* DNA required for successful hybrid capture, has not been measured.

Within-host *Mtb* genomic variation is of increasing interest clinically and epidemiologically. ^16–18^ Such within-host variation, sometimes referred to as minority variation when the *Mtb* population contains unfixed, or minority variants, which comprise less than 50% of the total population, can reflect a mixed or polyclonal infection of two distinct *Mtb* strains or it can reflect *de novo* variation generated through mutations occurring over the course of a single infection. As with fixed variants, the accuracy of minority variants identified via hybrid capture has not yet been evaluated. Previous studies have been inconclusive about the impact of hybrid capture on detection of minority variation.^8,19^ For example, one previous study found no difference between the minority variation detected in cultured and hybrid captured sputum.^5^

Our objective was to measure the efficiency and accuracy of hybrid capture as a tool for identification of both fixed and minority variants in *Mtb*. First, we conducted an artificial strain mixture experiment in which we serial diluted of DNA from well characterized *Mtb* isolates in human DNA, to simulate the overwhelmingly human DNA that is generated from clinical specimens. Second, we collected paired culture and sputa diagnostic samples from a national laboratory and compared the variation identified in paired samples.

## Methods

### Artificial mixture experiment using Mtb DNA standards

To measure the efficiency of hybrid capture and accuracy of variant identification in captured samples, we created mixtures of reference strains H37Ra (ATCC 25177) and X003899 (ATCC BAA-2237D-2) with well characterized genomes (representing *Mtb* lineages 4 and 2), in which the minor strain comprised 0, 1, 2, 5, 10, 20, and 50% of the input DNA (Fig. 1). We selected H37Ra and X003899 because they were well-characterized *Mtb* DNA isolates, both of which were genetically distinct from the commonly used reference genome, H37Rv. We then diluted the *Mtb* DNA with a human DNA standard (HTB-22D, ATCC HTB-22), so that total *Mtb* DNA comprised a small proportion of total input DNA by molecular weight (0.01%, 0.03%, 0.3%), approximating the proportion of *Mtb* DNA found in sputum with smear microscopy grades 1+, 2+, and 3+.^7^ For comparison, we also included a serial dilution that was 100% *Mtb* DNA.

**Figure 1.**
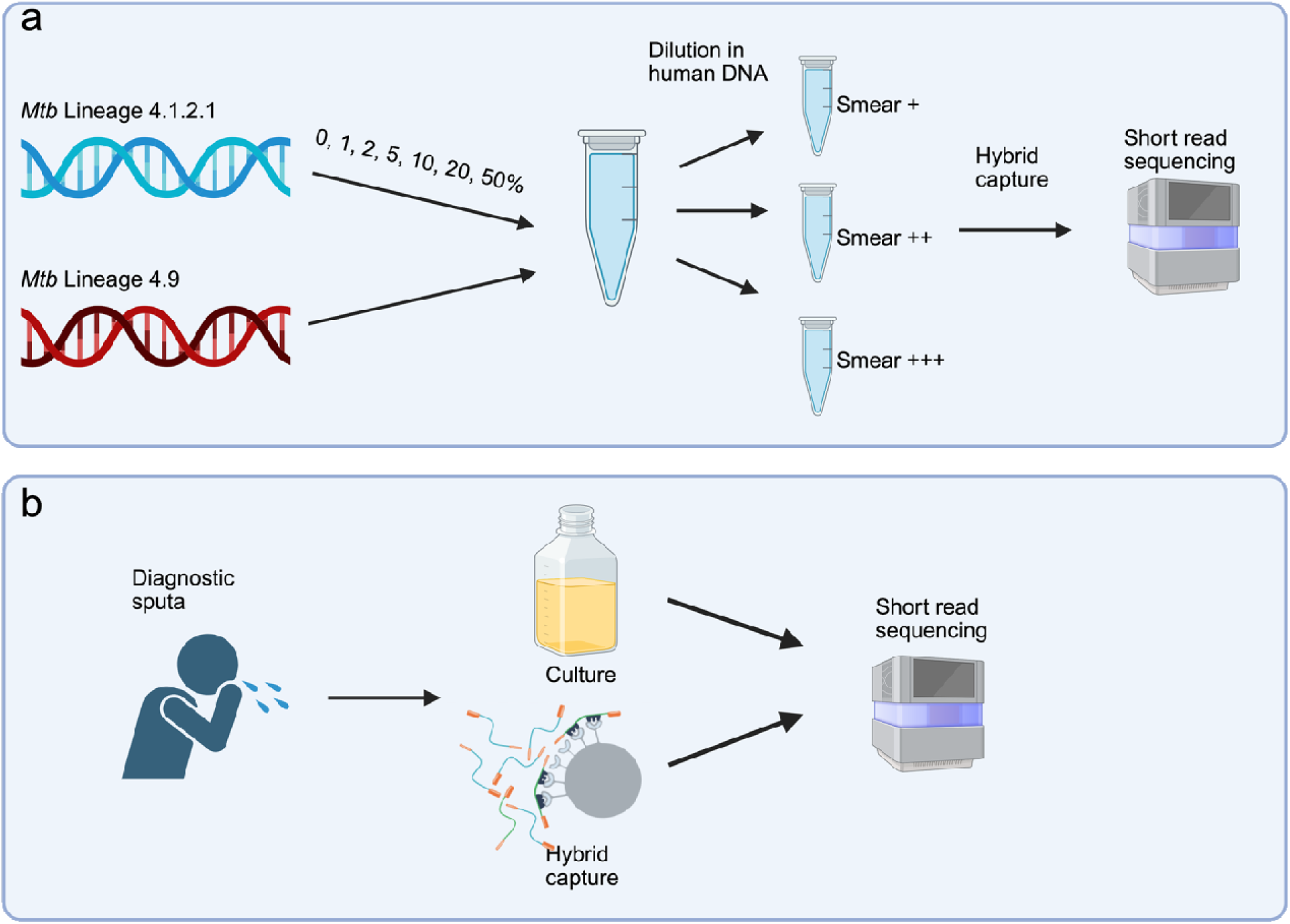
Experimental design. a) We conducted a serial dilution experiment in which we mixed two *Mtb* DNA standards representing lineage 4.1.2.1 (strain X003899) and 4.9 (strain H37ra), such that the minority strain represented 0-100% of the total DNA and then diluted *Mtb* DNA in human DNA, to simulate clinical samples which are overwhelmingly human DNA. We approximated the proportion of *Mtb* DNA found in sputum with smear microscopy grades 1+, 2+, and 3+.^7^ We used hybrid capture to enrich for *Mtb* DNA, conducted whole genome sequencing of captured libraries on an Illumina platform. b) We additionally prospectively sampled matched *Mtb* diagnostic cultures and unprocessed sputum submitted to ARUP diagnostic laboratories. We performed hybrid capture and Illumina whole genome sequencing on matched samples.

### Prospective sampling of paired sputum and cultured diagnostic samples

We additionally performed prospective genomic surveillance of paired sputa and diagnostic cultures at ARUP Laboratories, a national diagnostic laboratory in the United States (Fig. 1). We stored sputum samples in MTM solution and froze samples at -80°C until the time of processing. Prior to DNA extraction, 300 μL of decontaminated sputum samples were treated with 200 μL of a 2.5% saponin solution (Merck; diluted in 1D× PBS and sterile filtered). Samples were vortexed and incubated at room temperature for 10Dminutes to promote host cell lysis. 350 μL of nuclease-free water (Merck) was added for 30Dseconds after which 12 μL of 5 M NaCl (Merck) was added to the samples to create an osmotic shock. Samples were centrifuged at 6,000D×Dg for 5 min, and the pellets resuspended pellets in 100 μL 1D× PBS (Thermo Scientific).

15DμL DNase I [20,000 Units (50 to 375 U/μL); Thermo Scientific] and 11 μL of 10D× DNase buffer (Thermo Scientific) was added to digest free DNA. Samples were incubated in a thermomixer (Eppendorf, Hamburg, Germany) with shaking at 800 rpm at 37°C for 30 min. After incubation, samples were centrifuged at 6,000D×Dg for 3Dminutes, and the pellet washed twice with 800DμL 1D× PBS (Sriramulu et al., 2005). Finally, the pellet was resuspended in 300 μL of the lysis buffer of the extraction kit (Promega Maxwell RSC 48 Blood DNA kit) and extracted according to test manufacturer’s protocol.

We aliquoted 30 μL of DNA (target of ∼250 ng DNA required for sequencing).

For the paired diagnostic culture samples, prior to DNA extraction, 75 μL of the culture suspension was treated with 15 μL Proteinase K solution and incubated at room temperature for 15 minutes. 75 μL of lysis buffer was added and incubated at 95 °C for 30 minutes to kill viable mycobacterial cells. DNA from lysed samples were extracted according to manufacturer’s protocol.

### Ethics

We used de-identified remnant diagnostic samples and therefore the study was determined to be non-human subjects research by the University of Utah IRB (IRB 00176142).

### Genomic sequencing and variant identification

We prepared genomic libraries with the NEB Ultra II FS DNA Prep with myBaits Enrichment TB (Arbor Biosciences, D10008Myctb1). We conducted hybrid capture of *Mtb* using biotinylated RNA baits, tiled over the inferred *Mtb* ancestral genome,^20^ following the myBaits User Manual, Version 5.02. We pooled 8 libraries, 250 ng DNA each, for a total of 2 ug per capture reaction, as recommended for small genome capture in the myBaits User Manual, Version 5.02. We conducted paired-end sequencing on an Illumina NovaSeq X at the University of Utah High throughput Sequencing Core. Sequence data for all samples are available on the Sequence Read Archive (SRA), under SUB15840319.

We identified SNP variants with the *Mtb-call2* pipeline, as previously described.^21^ Briefly, the pipeline taxonomically filters reads with Kraken,^22^ maps reads with *bwa*^23^, and identifies genomic variants with *GATK* ^24^. Many *Mtb* genomic analysis pipelines routinely exclude the repetitive genes from the proline-glutamic acid and proline-proline-glutamic acid families, known as PE/PPE genes, as they are difficult for mapping algorithms and error-prone. We therefore stratified analysis by genomic regions outside and within the PE/PPE genes.

### Statistical analysis

Sequencing known mixtures of two *Mtb* DNA standards allowed us to use existing genomic resources to generate a “truth” VCF file of variants in H37Ra and X003899 with respect to the reference genome H37Rv. To do this, we used whole genomes sequences from H37Ra (GCF_001938725.1) and X003899 (supplied by ATCC). We used *MUMmer* to pairwise align each genome with the H37Rv genome and identified SNP variants using *nucmer*.^25^ We measured the performance (precision, recall, and F1 score) of variants identified in the serial dilution experiment using *hap.py,* a performance benchmarking tool for variant identification.^26^ We assessed the correlation between the expected minor allele frequency (proportion minority strain in the input mixture) and observed minor allele frequency for minority variants with Pearson’s correlation coefficient. We assigned true and false positive variants based on the majority allele in the input mixture.

For the artificial mixture samples and the diagnostic samples, we determined the percentage of reads taxonomically classified to the *Mycobacterium* genus by Kraken,^22^ percentage of reads mapping to *Mycobacterium tuberculosis* after filtering, and mean coverage with *Picard* CollectWgsMetrics.^27^ We additionally identified taxa outside of *Mycobacterium* with a large proportion of assigned sequence reads.

For the genomic surveillance samples, we quantified the number of minority variants as biallelic variants with a depth of 10X or greater for reference and alternative alleles. We measured the concordance of SNPs identified in cultured and captured samples with Jaccard’s index and measured the association between minor allele frequency for minority variants identified in both paired cultured and captured samples with Pearson’s correlation coefficient.

## Results

In the artificial strain mixture experiment, taxonomic assignment of reads to the *Mycobacterium* genus was high, even when *Mtb* comprised a small proportion of the input DNA (97% of reads were assigned to *Mycobacterium*, when *Mtb* DNA comprised 0.01% of the input) (Fig. 2a). The remaining assigned reads were largely human derived (mean: 0.3%, IQR: 2.6%). After taxonomic filtering, an average of 98% of *Mtb* reads mapped to the H37Rv reference genome (Fig. 2b). Taxonomic assignment and read-mapping efficiency after taxonomic filtering varied with the proportion of *Mtb* in the input sample as well as the genomic background (Fig. 2).

**Figure 2.**
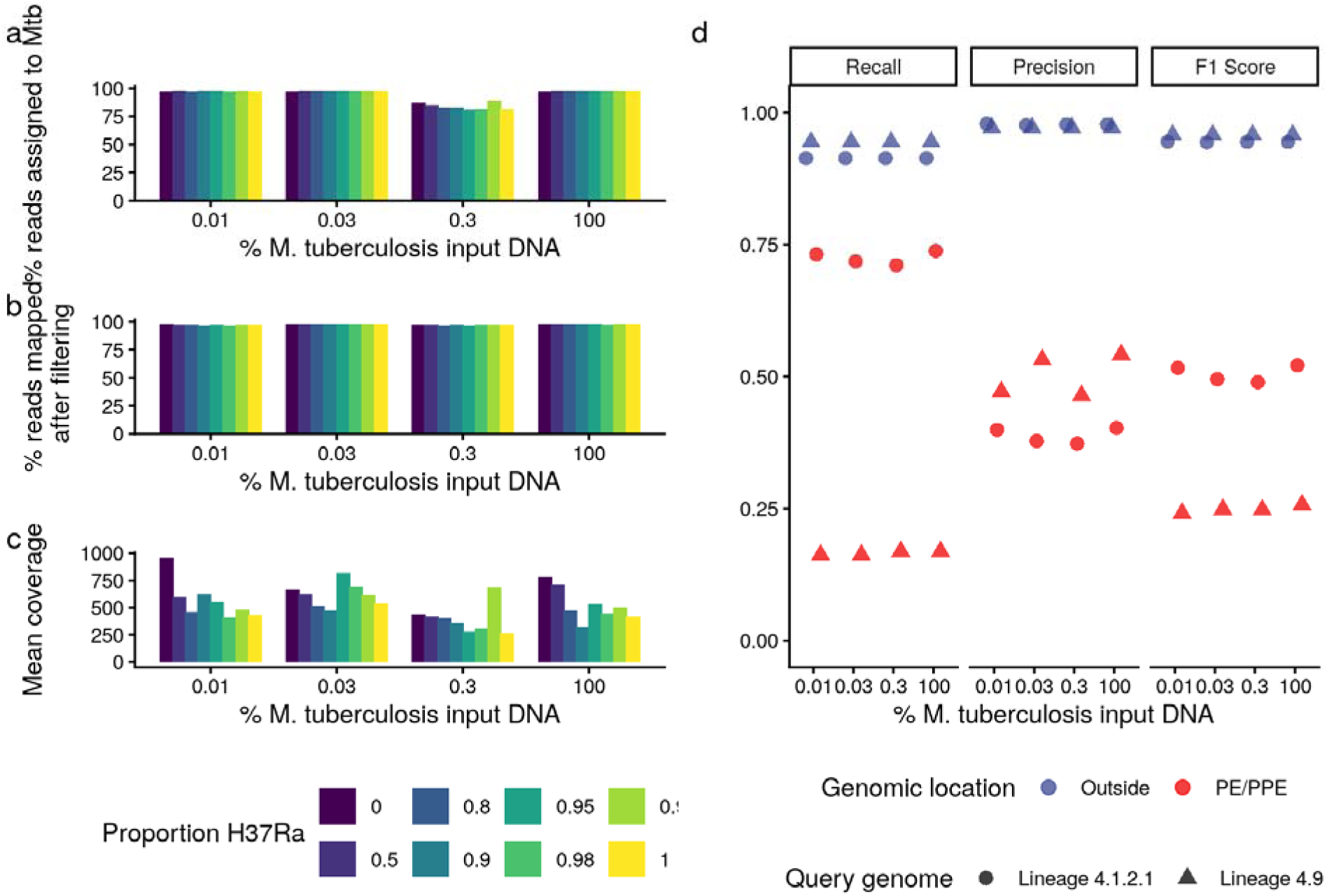
Performance of hybrid capture for experimental strain mixtures. (a) The percentage of reads taxonomically classified to the *Mycobacterium* genus by Kraken, (b) the percentage of reads mapping to *Mtb* after filtering, and mean coverage. The x-axis for both (a, b, and c) indicates the percentage input *Mtb* DNA in the total DNA mixture and color indicates the proportion H37Ra (color). (d) Precision and recall (sensitivity) of SNP identification, by percentage of *M. tuberculosis*, for the samples comprised of 100% Lineage 4.1.2.1 (points) and 100% 4.9 (triangles). Colors indicate genomic region: outside PE/PPE genes, blue; within PE/PPE genes, red.

Single nucleotide polymorphism (SNP) identification via hybrid capture had a mean recall of 0.91 and mean precision of 0.98 for the lineage 4.1.2.1 standard and a recall of 0.94 and precision of 0.97 for the lineage 4.9 standard, outside of PE/PPE genes (Fig. 2c). In PE/PPE genes, mean recall was 0.72 and precision of 0.39 for the lineage 4.1.2.1 standard, and recall was 0.17 and precision was 0.50 for the lineage 4.9 standard. Neither recall (p = 0.80) nor precision (p = 0.43) were impacted by the percentage of *Mtb* DNA in the input sample in separate binomial models.

Observed minor allele frequencies were closely correlated with expected minor allele frequencies (proportions of the minor strain) across dilutions (r = 0.60-0.79, p < 0.001) outside of PE/PPE genes (Fig. 3).

**Figure 3.**
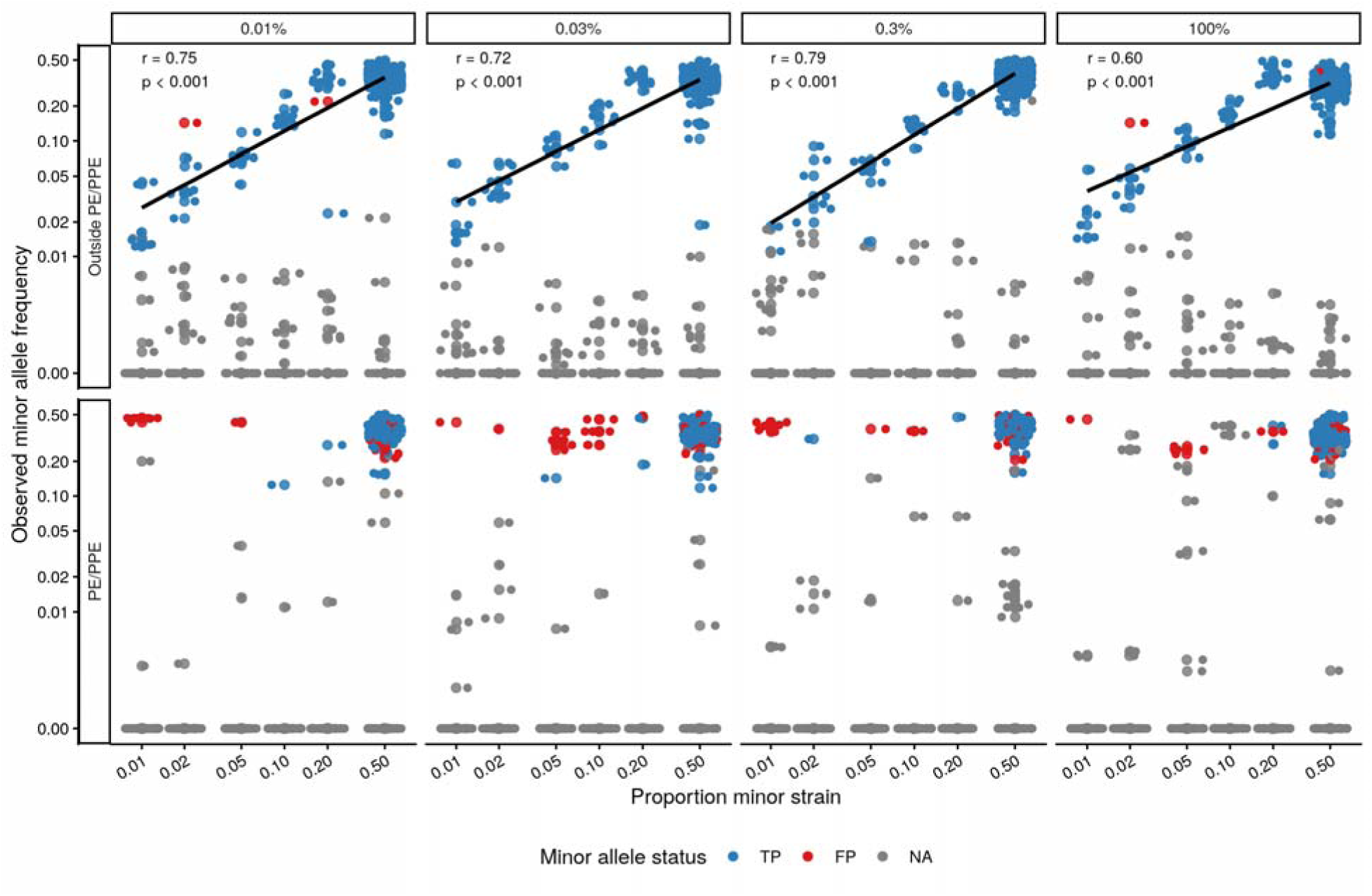
Proportion minor strain (expected minor allele frequency) versus observed minor allele frequency for minority variants identified, with columns indicating the percentage of *Mtb* DNA in the input DNA before hybrid capture and rows indicating genomic location. Colors indicate minor allele status.

Among clinical specimens, cultured samples had a higher percentage of reads assigned to *Mtb* (mean: 92%) than did paired captured samples (mean: 48%), prior to filtering. Four of seven captured samples had less than 75% of reads assigned to *Mtb* (Fig. 4a). We investigated taxonomic assignments for all captured samples and found that the majority of non-*Mtb* reads were assigned to the genus *Pseudomonas,* and largely, *Pseudomonas fluorescens*, a common environmental bacterium (Fig. 4b).

**Figure 4.**
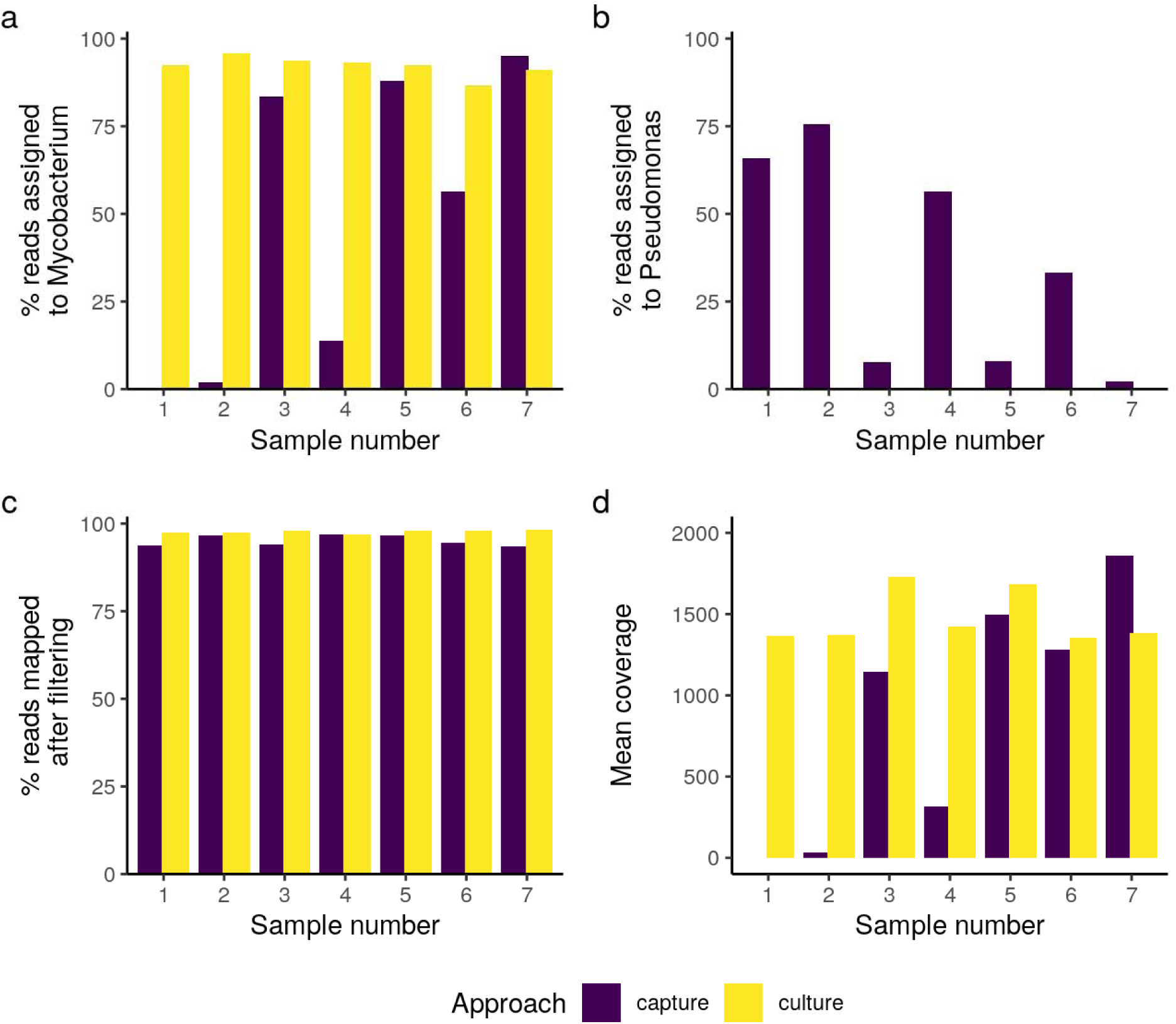
Performance of hybrid capture and culture for diagnostic sputa samples. For diagnostic sputa samples, a) the percentage of reads taxonomically classified to the *Mycobacterium* genus by Kraken for paired hybrid captured and cultured samples, b) the percentage of reads taxonomically classified to the *Pseudomonas* genus by Kraken, c) the percentage of reads mapping to *Mtb* after filtering, and d) mean coverage of the *Mtb* genome. Color indicates approach for *Mtb* enrichment: hybrid capture and culture.

After taxonomic filtering, the majority of reads for all captured samples mapped to *Mtb*, though mapping efficiency remained higher for cultured (98% reads) compared to captured samples (95% reads). Two samples had extremely low, <10X coverage, of the *Mtb* genome. Contamination levels were inversely correlated with coverage: mean coverage of the four samples that had >25% *Pseudomonas* contamination was 120X, compared to the mean coverage of 1980X for other samples, comparable to the depth of the cultured samples (1470X) (Fig. 4d).

We identified minority variants in each genomic surveillance sample with a minor allele frequency of at least 1% and minor allele depth of coverage of at least 5X (Fig. 5a). Few minority variants were consistently identified between cultured and captured samples, ranging from 0% (0 variants identified by both cultured and captured sample out of a total of 22 unique variants identified) to 13% (5/38 variants) outside PE and PPE genes (mean: 5%). Minority variants were more consistently identified by culture and capture in PE and PPE genes, ranging from 0% (0/49) to 45% (41/92) shared (mean: 18%). We did not find a difference between the number of minority variants detected in cultured (median: 14 variants, range: 9-23 variants) and captured samples (median: 22 variants, range 4-328 variants) (paired Wilcoxon test, p =0.2) outside of PE/PPE genes (Fig. 5b). Within PE/PPE genes, more minority variants were identified through culture (median: 70, range: 34-107) than hybrid captured samples (median: 36, range: 0-67, p = 0.03). Minor allele frequencies of minor variants identified in both culture and captured samples were correlated both outside (r = 0.95, p = 0.003) and within the PE/PPE genes (r = 0.63, p < 0.001) (Fig. 5c).

**Figure 5.**
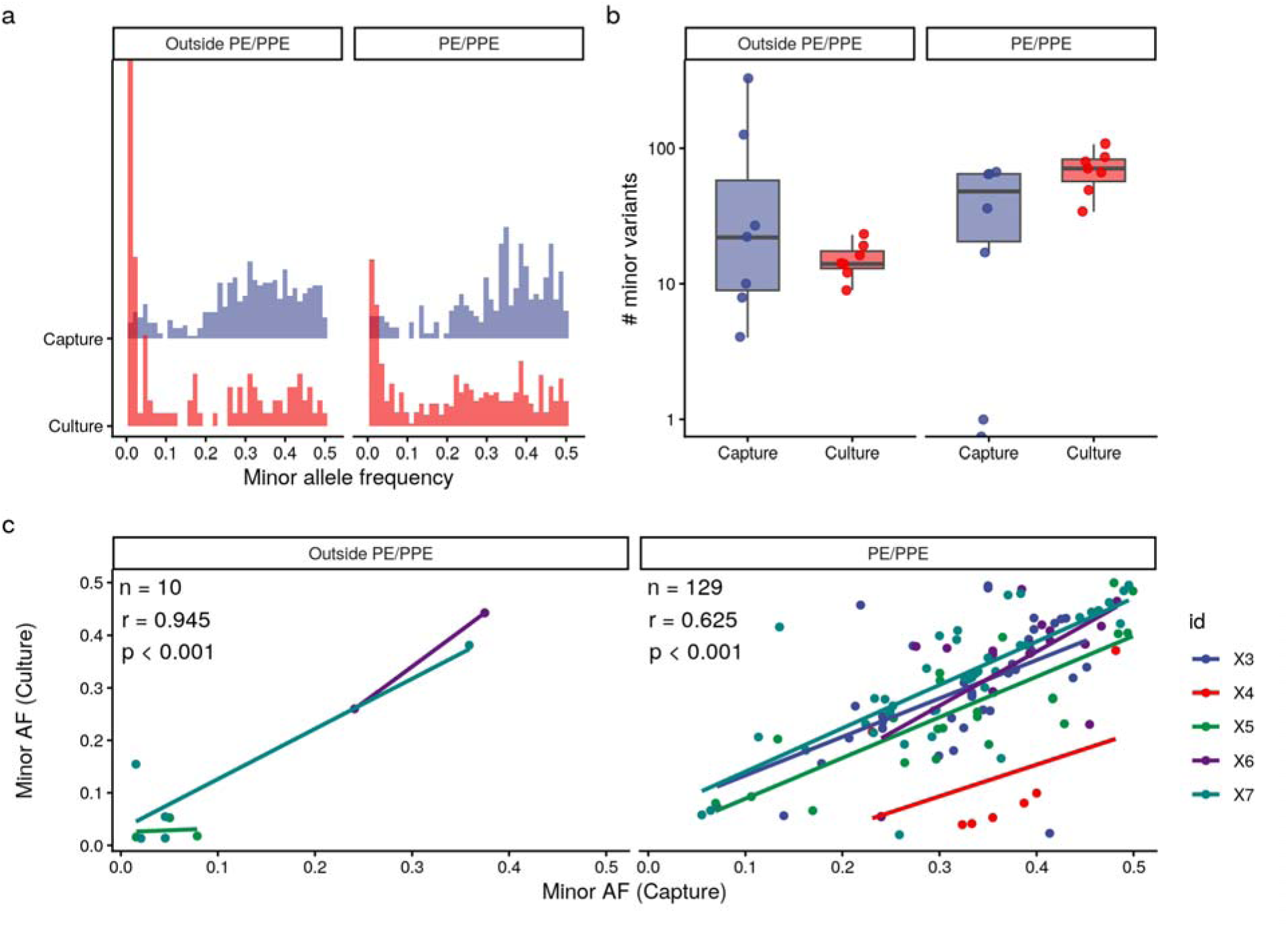
Minority variation recovered in paired clinical *Mtb* isolates. Below, we include all samples, regardless of *Pseudomonas* contamination. (a) Minor allele frequency distribution for cultured and captured samples. (b) The number of minor variants. Points indicate sample variant counts and box plots indicate the median, interquartile range, and whiskers extend to points within 1.5 times the IQR from the edges of the box. For visualization, the y-axis is on a log-scale. (c) Correlation of minor allele frequency between paired cultured and captured samples, with best fit lines drawn for each sample. Pearson’s correlation coefficient and p-value are reported by genomic location. No within-host variants were identified outside PE and PPE genes for sample X3.

Our findings were largely consistent when we excluded the four samples with a high degree of *Pseudomonas* contamination (>25% *Pseudomonas*) (Fig. S1). Few minority variants were consistently identified between cultured and captured samples, ranging from 0% (0 variants identified by both cultured and captured sample out of a total of 31 unique variants identified) to 13% (5/38 variants) outside PE and PPE genes (mean: 8% shared). Minority variants were more consistently identified by culture and capture in PE and PPE genes, ranging from 26% (37/98) to 45% (41/92) shared (mean: 36%). We did not find a difference between the number of minority variants detected in cultured (median: 15 variants, range: 9-19 variants) and captured samples (median: 22 variants, range 10-27 variants) (paired Wilcoxon test, p =0.42) outside of PE/PPE genes, nor within PE/PPE genes (p=0.42) (Fig S1). Minor allele frequencies of minor variants identified in both culture and captured samples were correlated both outside (r = 0.89, p = 0.003) and within the PE/PPE genes (r = 0.70, p < 0.001) (Fig. S1).

## Discussion

Genomic epidemiology is a powerful tool for understanding *Mtb* transmission dynamics and rates of antibiotic resistance but largely relies on mycobacterial culture, which is time consuming, requires specialized laboratory facilities, and may introduce a selective filter on the *Mtb* population. We evaluated hybrid capture as a potential replacement for culture. We found that hybrid capture enables efficient and accurate variant identification, even in samples overwhelmed by human DNA, and that it can be used to recover minority variation. Our findings indicate that hybrid capture is a promising tool for *Mtb* population genomic sequencing directly from diagnostic sputum samples that could expand the coverage of *Mtb* genomic epidemiology.

Previous studies have reported that hybrid capture can be used to sequence high coverage and *Mtb* genomes and generate genotypic resistance predictions concordant with phenotypic resistance, possibly more quickly than culture-based sequencing.^12,13^ Another earlier study employed hybrid capture to infer *Mtb* transmission linkages in Spain.^8^ Other studies have reported the recovery of minor variation in hybrid captured samples, but the effect of hybrid capture on the magnitude of recovered minority variation has been inconclusive. One study reported greater levels of within-host variation identified in captured samples compared to matched cultures,^19^ while another reported that minor variation identified by capture largely reflects what is present in culture.^8^ However, the efficiency of hybrid capture has not yet been measured across serial dilutions of *Mtb* DNA in human DNA, nor has hybrid capture been benchmarked for its performance in identifying both fixed or minority variants in DNA mixtures of well characterized isolates.

Our findings have important implications for genomic epidemiology studies relying on hybrid capture. Previous studies have demonstrated hybrid capture use can enable whole genome sequencing even in samples with low bacterial burden, including smear negative sputa.^8,19^ Consistent with these studies, we found that hybrid capture can be used for efficient sequencing, even when *Mtb* DNA is present at levels that reflect low bacterial burden, or smear grade 1+. As expected, variant identification was more accurate for the Lineage 4.9 isolate, than the Lineage 4.1.2.1 isolate, reflecting that capture probes were designed with the *H37Rv* reference genome, also in Lineage 4.9. This highlights a key bias in hybrid capture, that probes are based on previously characterized *Mtb* DNA, and will be less sensitive in genomes distant from the reference used in probe design. As expected, both recall and precision dropped in PE and PPE genes, known to be difficult to map, and often excluded from *Mtb* genomic analysis^28^. The higher recall of Lineage 4.1.2.1 compared to Lineage 4.9 variation in PE and PPE genes likely reflects the lower magnitude of true positive variation detected in pairwise alignment.

We find that bacterial contamination can overwhelm *Mtb* sequencing reads. While we do not know the source of the *Pseudomonas* contamination, it could have been introduced at any point of sputum sample processing or at the time of hybrid capture, since all samples were received and processed in the same diagnostic and sequencing laboratories. Taxonomic filtering is an important step both to exclude sequence reads that do not map to the *Mycobacterium* genus as well as to identify samples with high levels contamination that may need to be excluded from analysis.^29^ Yet taxonomic filtering is imperfect. In contaminated samples, we observed minority variation that persisted after filtering was concentrated in the *rrs* or 16S gene, which is highly conserved between *Mtb* and *Pseudomonas.* Excluding conserved microbial genes like *rrs* from variant identification in hybrid capture may be important. Further, even among samples with high levels of contamination, mapping efficiency, or the percentage of reads mapping to *Mtb* after taxonomic filtering, was high. Mapping efficiency thus is not a useful metric for identifying contamination, and sample-level filters that exclude samples with high levels of contamination are needed. For example, we detected high levels of minority variation in samples with high levels of *Pseudomonas* contamination, likely reflecting errors associated with incomplete taxonomic filtering of reads. However, our findings did not substantially change when excluding highly contaminated samples.

Our findings also support the use of hybrid capture for minor variant detection. In our serial dilution experiment, we found that input and observed minor allele frequencies were closely correlated in the serial dilution samples. Similarly, minor allele frequences were correlated between hybrid capture and cultured diagnostic sputa samples, evidence that hybrid capture can effectively capture two alleles at a genomic locus. The low levels of concordance between minor variants in capture and cultured samples may reflect true biological differences between the sample types, such as the previously described loss of variation in culture,^2^ or reflect biases in either approach, such as the impact of contaminating reads on variant detection. However, the underlying true pattern of variation for diagnostic samples is unknown and therefore, we cannot measure accuracy of minority variant identification. Input and observed minor allele frequencies as well as between captured and cultured samples are also correlated within the PE and PPE genes. Although false positive errors are also concentrated here, this highlights that excluding such variation likely excludes important biologically and epidemiologically meaningful signal.^30^

Our study has several limitations. First, we included only a small sample size of paired cultured and captured samples that were not contaminated. Studies with larger samples sizes are needed to establish the concordance of minority variants identified via culture and capture and to identify specific variant filters or mapping approaches that enable the recovery of high-quality minority variants from hybrid captured samples. Second, one of the limitations of hybrid capture and amplicon sequencing is that these approaches capture variation that is closely related to the reference genome used for probe or primer design. Such approaches may not be sensitive to large insertions or genomic rearrangements in samples with respect to the reference genome. Approaches that focus on host DNA depletion and metagenomic sequencing may be more effective in these cases. Third, probe synthesis for hybrid capture remains expensive, although some groups have reported decreasing per-sample costs.^8^ Optimizing cheaper alternatives and making *Mtb* probe design publicly available will increase the possibility that it can become a routine tool in population genomic sequencing and a viable tool for public health.

In conclusion, we find that hybrid capture efficiently enriches *Mtb* DNA from samples that are overwhelmingly human DNA and can accurately identify both consensus and minor variants. Hybrid capture offers an alternative to traditional culture-based sequencing workflows, enabling rapid genomic analysis without the need for mycobacterial culture facilities.

## Data Availability

Sequence data for all samples are available on the Sequence Read Archive (SRA), under SUB15840319.

## Acknowledgements

We would like to thank Megan Hirschi for sample processing and DNA extractions.

## Data Repository

Short Read Sequence Archive

## References

1. Meehan CJ, Goig GA, Kohl TA, et al. Whole genome sequencing of Mycobacterium tuberculosis: current standards and open issues. Nat Rev Microbiol. 2019;17(9):533–545. PMID:31209399

2. Mukamolova G V, Turapov O, Malkin J, Woltmann G, Barer MR. Resuscitation-promoting Factors Reveal an Occult Population of Tubercle Bacilli in Sputum.

3. Martín A, Herranz M, Ruiz Serrano MJ, Bouza E, García de Viedma D. The clonal composition of Mycobacterium tuberculosis in clinical specimens could be modified by culture. Tuberculosis (Edinb). 2010;90(3):201–207. PMID:20435520

4. Peter JG, Theron G, Muchinga TE, Govender U, Dheda K. The Diagnostic Accuracy of Urine-Based Xpert MTB/RIF in HIV-Infected Hospitalized Patients Who Are Smear-Negative or Sputum Scarce. PLoS One. 2012;7(7):e39966. PMID:22815718

5. Doughty EL, Sergeant MJ, Adetifa I, Antonio M, Pallen MJ. Culture-independent detection and characterisation of Mycobacterium tuberculosis and M. africanum in sputum samples using shotgun metagenomics on a benchtop sequencer. PeerJ. 2014;2:e585. PMID:25279265

6. Goig GA, Cancino-Muñoz I, Torres-Puente M, et al. Whole-genome sequencing of Mycobacterium tuberculosis directly from clinical samples for high-resolution genomic epidemiology and drug resistance surveillance: an observational study. Lancet Microbe. 2020;1(4):e175–e183.

7. Kok NA, Peker N, Schuele L, et al. Host DNA depletion can increase the sensitivity of Mycobacterium spp. detection through shotgun metagenomics in sputum. Front Microbiol. 2022;13:949328.

8. Charalampous T, Kay GL, Richardson H, et al. Nanopore metagenomics enables rapid clinical diagnosis of bacterial lower respiratory infection. Nature Biotechnology 2019 37:7. 2019;37(7):783-792. PMID:31235920

9. Votintseva AA, Bradley P, Pankhurst L, et al. Same-day diagnostic and surveillance data for tuberculosis via whole-genome sequencing of direct respiratory samples. J Clin Microbiol. 2017;55(5):1285–1298. PMID:17092190

10. Brown AC, Bryant JM, Einer-Jensen K, et al. Rapid whole-genome sequencing of mycobacterium tuberculosis isolates directly from clinical samples. J Clin Microbiol. 2015;53(7):2230–2237. PMID:25972414

11. Doyle RM, Burgess C, Williams R, et al. Direct Whole-Genome Sequencing of Sputum Accurately Identifies Drug-Resistant Mycobacterium tuberculosis Faster than MGIT Culture Sequencing. J Clin Microbiol. 2018;56(8):666–684. PMID:29848567

12. Kalinich CC, Gonzalez FL, Osmaston A, et al. Tiled Amplicon Sequencing Enables Culture-free Whole-Genome Sequencing of Pathogenic Bacteria From Clinical Specimens. bioRxiv. Published online December 20, 2024:2024.12.19.629550. PMID:39763738

13. Verma R, Moreira FMF, do Prado Morais AO, Walter KS, Santos PCP Dos, … Detection of M. tuberculosis in the environment as a tool for identifying high-risk locations for tuberculosis transmission. Science of the Total Environment. 2022;(843, 156970).

14. Cohen T, van Helden PD, Wilson D, et al. Mixed-Strain Mycobacterium tuberculosis Infections and the Implications for Tuberculosis Treatment and Control. Clin Microbiol Rev. 2012;25(4):708–719. PMID:23034327

15. Sobkowiak B, Cudahy P, Chitwood MH, et al. A new method for detecting mixed Mycobacterium tuberculosis infection and reconstructing constituent strains provides insights into transmission. Genome Med. 2025;17(1):8.

16. Walter KS, Cohen T, Mathema B, et al. Signatures of transmission in within-host Mycobacterium tuberculosis complex variation: a retrospective genomic epidemiology study. Lancet Microbe. Published online 2024.

17. Nimmo C, Shaw LP, Doyle R, et al. Whole genome sequencing Mycobacterium tuberculosis directly from sputum identifies more genetic diversity than sequencing from culture. BMC Genomics. 2019;20(1):1–9. PMID:31109296

18. Comas Ĩ, Chakravartti J, Small PM, et al. Human T cell epitopes of Mycobacterium tuberculosis are evolutionarily hyperconserved. Nat Genet. 2010;42(6):498–503. PMID:20495566

19. Walter KS, Santos PCP Dos, Gonçalves TO, da Silva BO, … The role of prisons in disseminating tuberculosis in Brazil: a genomic epidemiology study. The Lancet Regional Health–Americas. 2022;(9).

20. Wood DE, Salzberg SL. Kraken: Ultrafast metagenomic sequence classification using exact alignments. Genome Biol. 2014;15(3):R46. PMID:24580807

21. Li H, Durbin R. Fast and accurate short read alignment with Burrows-Wheeler transform. Bioinformatics. 2009;25(14):1754–1760. PMID:19451168

22. Walter KS, Colijn C, Cohen T, et al. Genomic variant-identification methods may alter Mycobacterium tuberculosis transmission inferences. Microb Genom. 2020;6.

23. Kurtz S, Phillippy A, Delcher AL, et al. Versatile and open software for comparing large genomes. Genome Biol. 2004;5(2):R12. PMID:14759262

24. Krusche P. Haplotype Comparison Tools, happy. Published online 2021.

25. Picard Tools - By Broad Institute.

26. Ates LS. New insights into the mycobacterial PE and PPE proteins provide a framework for future research. Mol Microbiol. Published online 2019:0–2. PMID:31661176

27. Goig GA, Blanco S, Garcia-Basteiro AL, Comas I. Contaminant DNA in bacterial sequencing experiments is a major source of false genetic variability. BMC Biol. 2020;18(1). PMID:32122347

28. Walter KS, Colijn C, Cohen T, et al. Genomic variant-identification methods may alter Mycobacterium tuberculosis transmission inferences. Microb Genom. 2020;(6 (8)).

