## Supplementary Information for "Optimizing culture-free approaches to recover high quality *M. tuberculosis* genomic variation"

**Supplementary Figure 1. Minority variation recovered in paired clinical *Mtb* isolates.** We include variants with a minor allele frequency of 1% and with minor allele support of at least 5X. Below, we include the three paired samples with less than 25% *Pseudomonas* contamination; Figure 5 includes all samples. (a) Minor allele frequency distribution for cultured and captured samples. (b) The number of minor variants. Points indicate sample variant counts and box plots indicate the median, interquartile range, and whiskers extend to points within 1.5 times the IQR from the edges of the box. (c) Correlation of minor allele frequency between paired cultured and captured samples, with best fit lines drawn for each sample. Pearson’s correlation coefficient and p-value are reported by genomic location. No within-host variants were identified outside PE and PPE genes for sample X3.

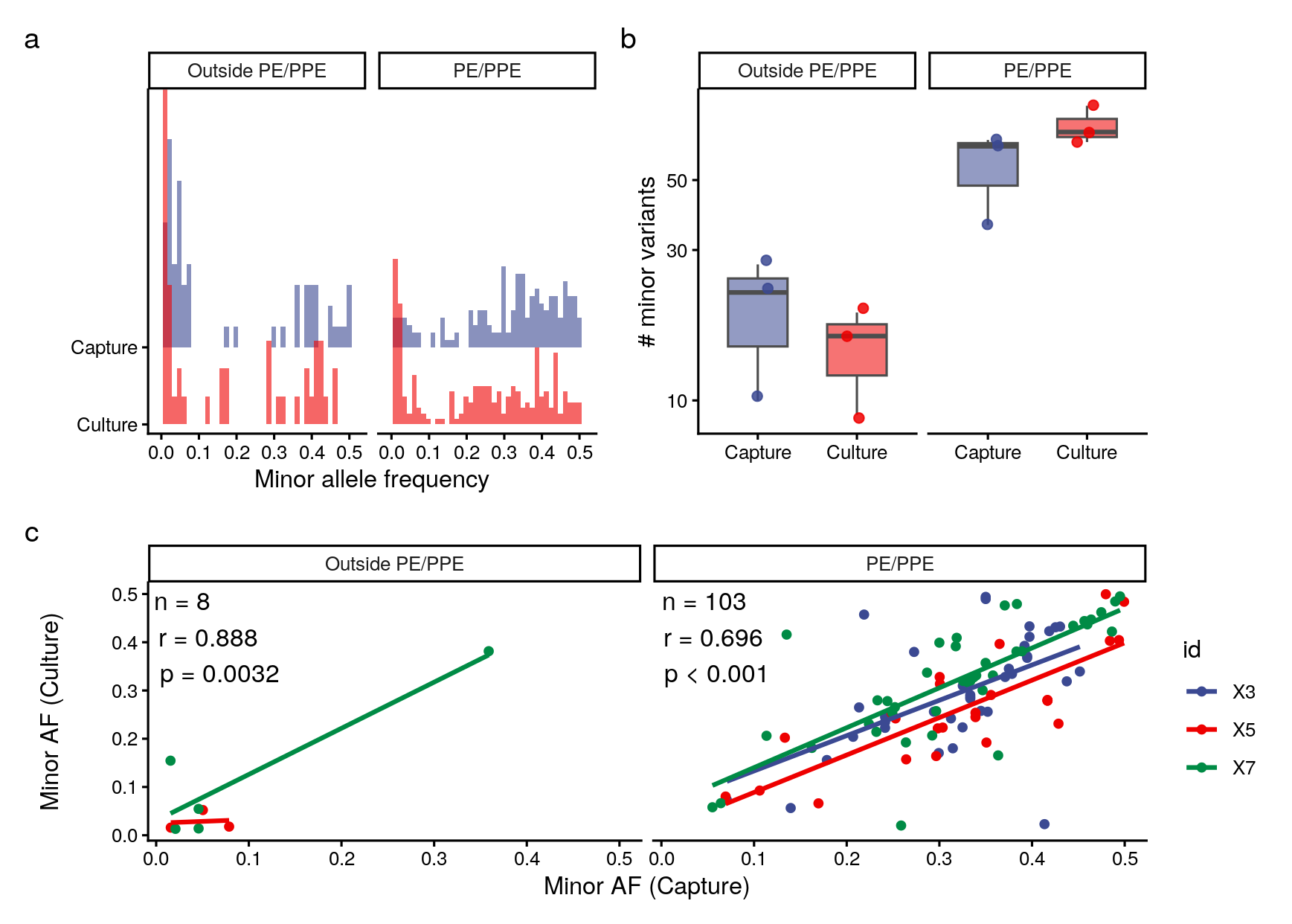
